# Stage-Aware and Explainable Machine Learning for Predicting IVF Implantation

**DOI:** 10.64898/2026.10.13.26365427

**Authors:** Harshil Sharma, Kevine Yemelong, Ana Carmona, Zhila Naghibi Harat, Bahareh Rahmani

**Author notes:** Corresponding Author: Dr. Bahareh Rahmani.

## Abstract

Predicting implantation outcomes in vitro fertilization (IVF) is challenging because clinically relevant information becomes available at different points during treatment. In this study, we developed a stage-aware, leakage-conscious, and explainable machine-learning framework to predict implantation using a public IVF dataset. β-HCG was excluded to avoid potential downstream target leakage. The data were divided into training, validation, and independent test sets. Several machine-learning models, including Logistic Regression, Random Forest (RF), Extra Trees, XGBoost, and Hist Gradient Boosting, were optimized using stratified cross-validation. A fixed RF model was then evaluated at three stages of the IVF process, using baseline couple characteristics, gamete and fertilization variables, and embryological variables, respectively. Model performance improved as additional treatment-related information became available, with the largest improvement observed after the inclusion of embryological variables. The final RF model showed good predictive performance on the independent test set. The number of embryos was consistently identified as the most important predictor across RF, permutation, and SHAP analyses. Overall, the findings suggest that IVF implantation prediction becomes more accurate as later-stage treatment information is incorporated, particularly information related to embryological characteristics. The proposed approach also provides interpretable predictions while reducing the risk of information leakage. Further evaluation using independent clinical datasets is needed before the framework can be considered for clinical use.

## 1. Introduction

In vitro fertilization (IVF) is a complex and stepwise treatment process. Its success can be influenced by many factors, including couple characteristics, ovarian reserve, gamete quality, fertilization outcomes, embryo development, and factors related to embryo transfer. Because infertility affects many individuals and couples worldwide, IVF has become an important part of infertility treatment [1]. At the same time, IVF generates a large amount of clinical and laboratory information, creating opportunities to use artificial intelligence (AI) and machine-learning methods to support treatment and clinical decision-making [2,3].

AI has been increasingly used in different areas of assisted reproductive technology. In the IVF laboratory, AI-based approaches have been explored for analyzing clinical and laboratory information and supporting different stages of treatment [2]. AI has also been proposed as a tool for more personalized fertility treatment, including treatment planning and outcome prediction [3]. As these models become more common in healthcare, understanding how they arrive at their predictions has also become increasingly important. Explainable AI (XAI) methods can help researchers and clinicians examine how individual variables contribute to model predictions [4,5].

Machine-learning methods are particularly useful for clinical datasets that contain multiple variables and potentially complex relationships. Random Forest, for example, combines multiple decision trees and can capture nonlinear relationships between predictors and outcomes [6]. XGBoost is another widely used tree-based method that can model complex patterns in structured data [7]. Other computational approaches, including genetic algorithms, statistical pattern-recognition methods, and deep learning, have also been widely used in predictive modeling and AI [8–10]. Software libraries such as scikit-learn and PyTorch have made these methods more accessible for developing and evaluating machine-learning models [11,12].

AI and machine learning have also been applied to different stages of IVF, including treatment-related decision-making and embryo assessment. By combining information from multiple sources, these methods may help estimate the likelihood of treatment outcomes and provide additional information for clinical decision-making. However, a model that performs well statistically is not necessarily ready for clinical use. Prediction models should also provide reliable probability estimates and should be evaluated using appropriate validation methods [13]. Transparent reporting of model development, evaluation, and validation is important for assessing the reliability and reproducibility of clinical prediction models [14].

Another important consideration is understanding which variables are driving a model’s predictions. Explainability methods such as SHAP (SHapley Additive exPlanations) can estimate the contribution of individual features to model predictions and help identify the variables that have the greatest influence on the model [15]. This information can be particularly useful in healthcare because it allows researchers to examine whether the model is relying on clinically meaningful information rather than simply treating the model as a black box.

An additional challenge in IVF prediction is that not all clinical information is available at the same time. Some characteristics, such as couple demographics and baseline clinical information, are available before treatment begins. Information about gametes and fertilization becomes available later, while detailed embryological information is available only after subsequent laboratory procedures. Therefore, a prediction model that uses all available information may provide a good overall prediction but may not reflect what could realistically have been predicted at an earlier stage of treatment.

The timing of information is also closely related to the problem of data leakage. Including information that becomes available after the prediction point, or information that is closely related to the outcome itself, can make a model appear more accurate than it would be in real-world use. For this reason, predictors should be considered according to when they become available during the IVF process. In this study, β-HCG was excluded because it represents downstream information that could contribute to target leakage.

To address these issues, we evaluated the same machine-learning model at three sequential stages of the IVF process. The first stage included baseline characteristics, the second incorporated gamete and fertilization information, and the third included embryological variables. This approach allowed us to examine how predictive performance changed as additional information became available during treatment, rather than combining all variables into a single model without considering their timing.

The aim of this study was to develop and evaluate a stage-aware, leakage-conscious, and explainable machine-learning framework for IVF implantation prediction. We specifically examined the additional predictive value gained when gamete, fertilization, and embryological information became available during treatment. We also used feature-importance and SHAP analyses to better understand which variables contributed most strongly to the model’s predictions. By considering both the timing and interpretability of the available information, this study seeks to provide a practical framework for evaluating machine-learning approaches to IVF implantation prediction.

## 2. Data Description

The analysis used the public Comprehensive Dataset Profile of In Vitro Fertilization (IVF) Clinical and Embryological Data [28]. Of 5,002 records, two lacked a valid target, leaving 5,000 observations (2,508 failed and 2,492 successful implantations). Eighteen predictors were retained: female and male age, number of attempts, AMH, Medicines and treatments, oocyte maturity measures (MII, MI, other), sperm count, motility and morphology, number of fertilized eggs, Embryo Grades 1-4, Number of embryos, and the source variable frezzing. β-HCG was excluded because it is downstream of transfer and closely aligned with pregnancy establishment; Egg retrieval date was also excluded. Female age ranged from 20 to 49 years.

## 3. Methodology

Records were stratified into training (70%), validation (15%), and independent test (15%) partitions (random state 42). Logistic Regression, RF, Extra Trees, XGBoost, and HistGradientBoosting were tuned on training data using randomized search with five-fold stratified cross-validation and ROC-AUC as the objective. Validation-set thresholds were selected using Youden’s J statistic. RF and XGBoost were included as established nonlinear ensemble methods [20] [21].

For the stage-wise analysis, the same RF configuration was retained across stages (300 trees, minimum leaf size 8, max_features=0.5, balanced class weighting). Stage 1 used eight baseline/couple predictors; Stage 2 added oocyte maturity and fertilization variables; Stage 3 added embryo grades, number of embryos, and frezzing. Paired bootstrap resampling (2,000 iterations) quantified AUC gains, while final metric uncertainty used 3,000 bootstrap resamples. RF feature importance, permutation importance, and SHAP [19] were used for interpretation. Calibration was evaluated with Brier score, intercept, and slope, and decision-curve analysis followed Vickers and Elkin [22].

## 4. Performance Evaluation

ROC-AUC was the primary discrimination metric. Accuracy, balanced accuracy, precision, sensitivity, specificity, F1 score, average precision, Matthews correlation coefficient, and Brier score were also calculated. A calibration intercept near 0 and slope near 1 were considered favorable [17]. Table 1 shows the validation and reporting followed general prediction-model principles [23] [24] [25].

**Table 1.** Independent test performance of candidate models.

| Model | AUC | Acc. | Sens. | Spec. | F1 | Brier |
| --- | --- | --- | --- | --- | --- | --- |
| Random Forest | 0.894 | 0.805 | 0.877 | 0.734 | 0.818 | 0.131 |
| Extra Trees | 0.892 | 0.785 | 0.730 | 0.840 | 0.772 | 0.132 |
| HistGradientBoosting | 0.889 | 0.797 | 0.781 | 0.814 | 0.793 | 0.133 |
| XGBoost | 0.888 | 0.803 | 0.789 | 0.816 | 0.799 | 0.133 |
| Logistic Regression | 0.841 | 0.765 | 0.826 | 0.705 | 0.778 | 0.165 |

## 5. Results

The nonlinear ensembles showed similar test discrimination. RF was numerically highest (AUC 0.8938), followed by Extra Trees (0.8923), HistGradientBoosting (0.8888), XGBoost (0.8882), and Logistic Regression (0.8405); the small differences among the four nonlinear models do not establish statistical superiority (figure 1).

**Figure 1.**
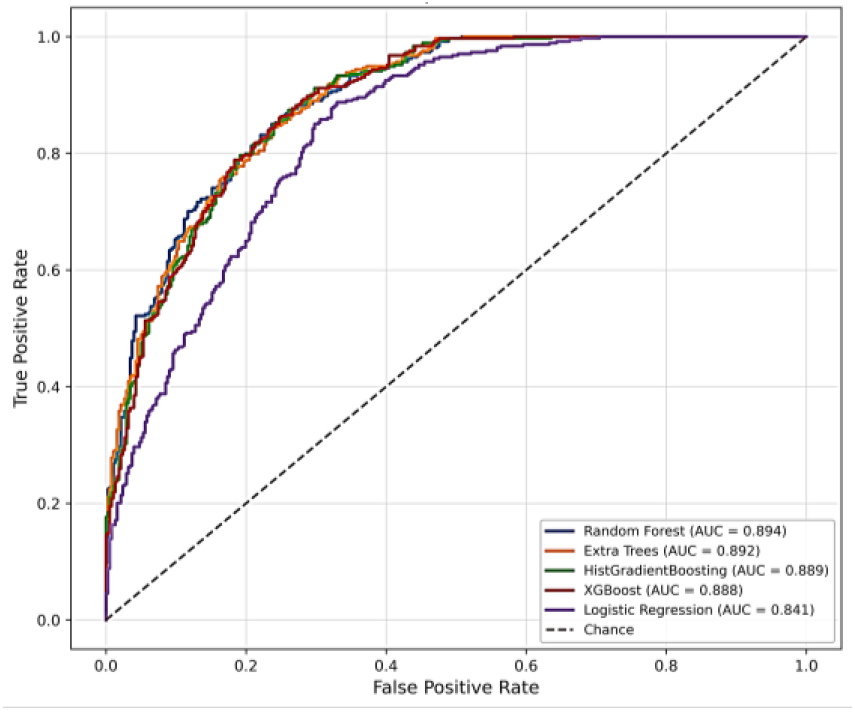
Independent test ROC curves for the five candidate algorithms.

Stage-wise performance increased progressively. Stage 1 AUC was 0.7378, Stage 2 AUC was 0.8033, and Stage 3 AUC was 0.8951. The gains were 0.0655 for Stage 1→2 (95% CI 0.0361-0.0979), 0.0918 for Stage 2→3 (95% CI 0.0664-0.1172), and 0.1573 overall (95% CI 0.1271-0.1900) (Table 2) (Figure 2).

**Table 2.**
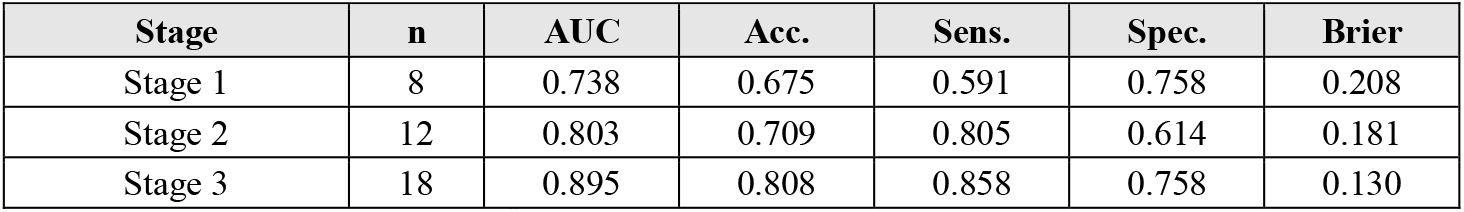
Stage-wise Random Forest performance.

| Stage | n | AUC | Acc. | Sens. | Spec. | Brier |
| --- | --- | --- | --- | --- | --- | --- |
| Stage 1 | 8 | 0.738 | 0.675 | 0.591 | 0.758 | 0.208 |
| Stage 2 | 12 | 0.803 | 0.709 | 0.805 | 0.614 | 0.181 |
| Stage 3 | 18 | 0.895 | 0.808 | 0.858 | 0.758 | 0.130 |

**Figure 2.**
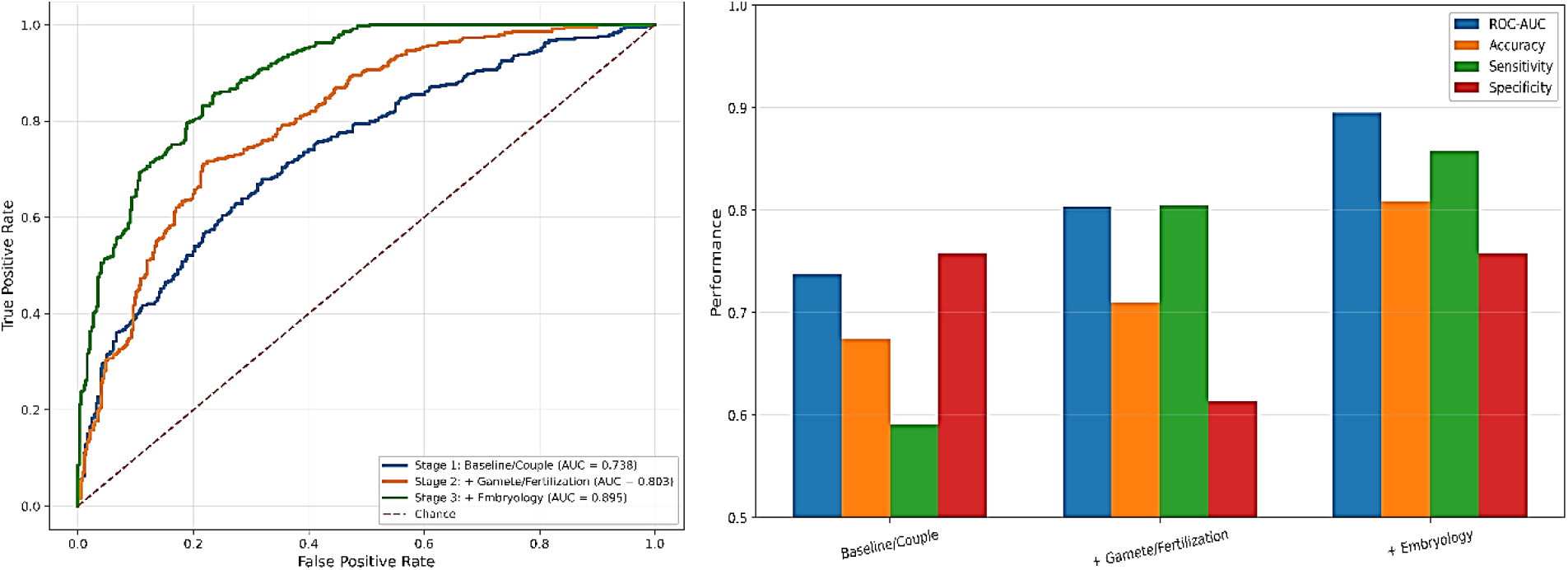
(A) Stage-wise ROC curves and (B) performance across sequential IVF information stages.

Figure 3 shows bootstrap evaluation of the final Stage 3 RF yielded AUC 0.8955 (95% CI 0.8732-0.9161), accuracy 0.8083, sensitivity 0.8585, specifically 0.7585, F1 0.8168, MCC 0.6200, and Brier score 0.1302. Number of embryos ranked first by RF importance (0.4251), permutation importance (0.2319), and mean absolute SHAP (0.2134), followed by freezing, Medicines and treatments, fertilized eggs, and Embryo Grade 1.

**Figure 3.**
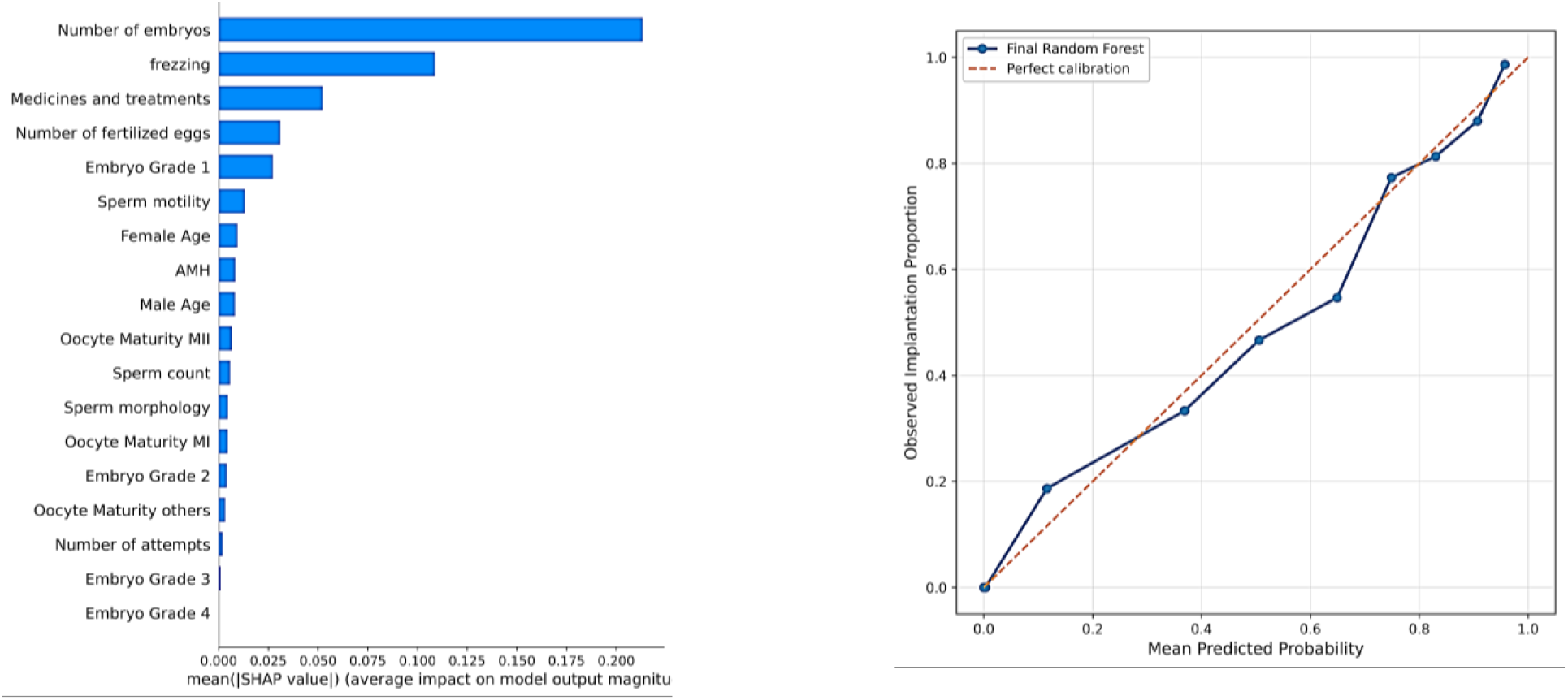
(A) Global SHAP importance for the final Stage 3 Random Forest and (B) calibration curve (intercept −0.0690; slope 0.9807).

**Figure 4.**
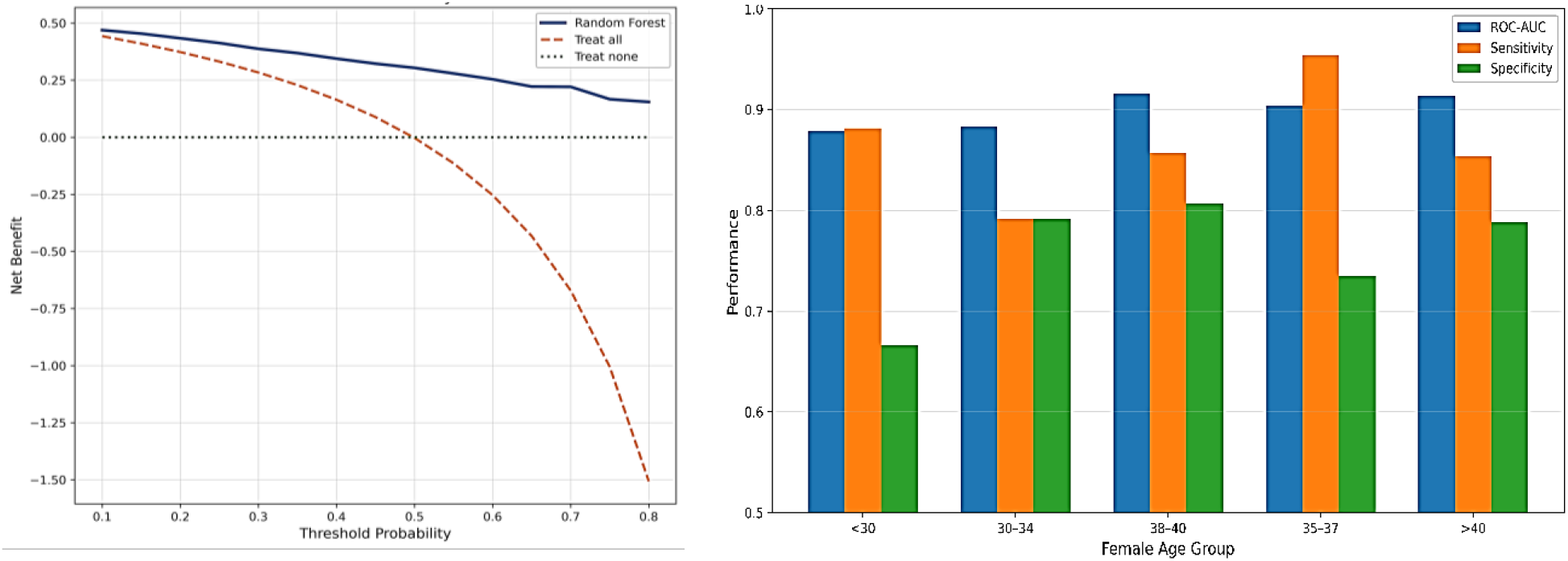
(A) Age-stratified performance and (B) exploration decision-curve analysis over threshold probabilities 0.10-0.80.

Age-stratified AUCs were 0.8784 (<30 years), 0.8828 (30-34), 0.9038 (35-37), 0.9153 (38-40), and 0.9139 (>40), indicating broadly stable discrimination. Exploratory decision-curve analysis showed higher estimated net benefits than treat-all and treat-none strategies across the evaluated thresholds.

## 6. Discussion

The main finding is that predictive discrimination increased as later-stage IVF information became available. The largest incremental gain occurred after embryological variables were incorporated, consistent with the sequential nature of IVF and prior adaptive prediction work [8]. The final AUC was also consistent with literature showing useful performance of nonlinear ML approaches for IVF outcomes [5] [6] [7] [9] [10].

Explainability supported the stage-wise result: embryo-related variables, particularly Number of embryos, dominated model attribution. Prior studies likewise show predictive value in embryo characteristics and embryo-focused AI systems [11] [13] [14] [15]. SHAP and feature importance, however, describe model dependence and should not be interpreted as causal effects [19]. Female age remained predictive but was less influential than several embryo-stage features; performance nevertheless remained stable across the examined age strata.

Calibration was favorable in the held-out sample, with slope 0.9807 and intercept −.0690, but internal calibration and DCA do not establish transportability or prospective clinical benefit [17] [22]. Important limitations include secondary public data, incomplete clinical definitions for frezzing and Medicines and treatments, inability to confirm whether repeated cycles from the same patient were present, and absence of external validation. External and preferably prospective multicenter validation is therefore required [23] [24] [25]. Overall, the results support a dynamic, stage-dependent IVF prediction framework in which probabilities are updated as new treatment information becomes available.

## Data Availability

Maghdid S, Rashid TA, Askar S. Comprehensive Dataset Profile of In Vitro Fertilization (IVF) Clinical and Embryological Data. Mendeley Data. 2026. Version 1. doi:10.17632/hd8yy6gzc5.1. [Dataset source; not part of the pre-2025 research literature.]

https://data.mendeley.com/datasets/hd8yy6gzc5/2

## Declarations

This study used a publicly available secondary dataset and involved no direct participant recruitment or intervention. The authors claim no ethics approval for the original data collection; original ethics and consent information should be reported from authoritative dataset documentation. Consent for publication: not applicable. Competing interests: none declared. Funding: no specific funding. Data availability: Mendeley Data [28]. Harshil Sharma contributed to conceptualization, methodology, software, formal analysis, visualization, interpretation, and manuscript preparation. Large Language Model (LLM)-based tools were used only for language refinement and manuscript organization; all scientific content, analysis, interpretation, and conclusions were independently verified by the authors.

